# Development and reliability and validity test of the Questionnaire on Knowledge, Attitude and Practice of ICU Nurses on Blood Oxygen Saturation Management in Mechanically Ventilated Patients

**DOI:** 10.64898/2026.06.13.26355567

**Authors:** Qian Xie, Hong Bian, Ping Yu, Ping He

## Abstract

**Objective:** A questionnaire on the knowledge, attitude and practice of ICU nurses regarding the management of blood oxygen saturation in patients with mechanical ventilation was compiled, and its reliability and validity were tested.

**Method:** Drawing upon the knowledge-attitude-practice theory, the initial questionnaire draft was developed through literature review and consultation with Delphi experts. Employing convenience sampling, 32 nurses from the General ICU of Wuxi Second People’s Hospital were surveyed between 1 August 2025 and 27 September 2025, enabling item screening and assessment of reliability and validity.The full version of the developed questionnaire is provided as Supporting Information (S1 File). All items are published under a CC BY 4.0 license, which permits unrestricted use, distribution, and reproduction in any medium, provided the original author and source are credited.

**Result:** A questionnaire on the knowledge, attitude and practice of ICU nurses regarding the management of blood oxygen saturation in mechanically ventilated patients was finalised, comprising 26 items: 11 in the knowledge dimension, 6 in the attitude dimension and 9 in the behaviour dimension. The overall Cronbach’s α coefficient for the questionnaire was 0.88, with dimension-specific coefficients of 0.787, 0.722, and 0.781 respectively. The Spearman-Brown coefficient for the entire questionnaire was 0.967, while dimension-specific coefficients were 0.796, 0.666, and 0.728 respectively. The content validity index at the questionnaire level (S-CVI) was 0.886, and the item-level content validity index (I-CVI) ranged from 0.913 to 0.967. 0.728. The questionnaire’s level content validity index (S-CVI) was 0.886, and the item level content validity index (I-CVI) ranged from 0.913 to 1.00.

**Conclusion:** The questionnaire on knowledge, attitude and practice of blood oxygen saturation management in mechanically ventilated patients demonstrates good reliability and validity. It may serve as an assessment tool for intensive care unit nurses regarding their knowledge, attitude, and practices concerning blood oxygen saturation management in mechanically ventilated patients, thereby establishing a foundation for developing targeted intervention strategies in future practice.

## Introduction

Patients in intensive care units (ICUs) frequently present with severe multi-organ dysfunction, with respiratory failure being a common and life-threatening complication ^[1]^. Mechanical ventilation (MV) is a critical life-sustaining technique for ICU patients ^[2][3]^.The most readily accessible vital sign reflecting a patient’s oxygenation status is their blood oxygen saturation (Sp). Accurate monitoring and effective management of SpO_2_ constitute a critical component of ICU nursing practice. InadequateSpO_2_management, whether manifesting as hypoxaemia or hyperoxaemia, may lead to adverse outcomes such as organ dysfunction and increased infection risk ^[2]^.As primary caregivers for mechanically ventilated patients, ICU nurses bear crucial responsibilities including vital sign monitoring, ventilator parameter adjustment, and oxygen therapy administration. Their knowledge, attitude, and practices (KAP) regarding SpO_2_ management constitute core determinants of management quality.Currently, no dedicated questionnaire exists internationally or domestically for assessing ICU nurses’ SpO_2_management of_mechanically_ ventilated patients. Existing evaluation methods remain fragmented, lacking systematic rigour and specificity, and thus fail to comprehensively and accurately gauge nurses’ knowledge, attitude, and practices in this domain. This impedes precise identification of deficiencies in SpO_2_ management, thereby undermining the development of standardised training protocols.The Knowledge-Attitude-Behaviour (KAB) theory, first proposed by Koster in 1976, is a theoretical model of human health behaviour involving three sequential processes: acquiring knowledge, altering attitude, and forming behaviour ^[4]^. This theoretical framework has been extensively applied in health behaviour management programmes across public health disease prevention, medical care, and nursing domains (^[4,5])^.Therefore, grounded in the KSE framework and integrated with ICU clinical practice, a questionnaire specifically designed for ICU nurses to manage SpO_2_ in mechanically ventilated patients was developed. This questionnaire comprehensively addresses all aspects of SpO_2_ management and accurately reflects nurses’ knowledge, attitude, and practices regarding SpO_2_ management in mechanically ventilated patients, providing a reliable tool for clinical application and management.

## 1 Research Methodology

### 1.1 Initial Questionnaire Development

Between 17 May and 28 June 2025, the research team comprehensively reviewed the latest domestic and international literature ^[6-19]^on mechanical ventilation, SpO_2_ management, ICU nursing, and nursing knowledge-attitude-behaviour questionnaires, grounded in the KBE theoretical model.Particular emphasis was placed on relevant clinical guidelines, expert consensus statements, and nursing practice standards to identify key knowledge points, nursing principles, and practical considerations for SpO_2_ management in mechanically ventilated patients. Initial questionnaire items were developed by integrating the Clinical Practice Guidelines for Mechanical Ventilation and the competency requirements for ICU nursing roles. The initial version comprised 26 items across three dimensions: knowledge (11 items), attitude (6 items), and behaviour (9 items).

### 1.2 Expert Consultation

Expert inclusion criteria: (1) Bachelor’s degree or higher; (2) Intermediate-level professional title or above; (3) Experience in nursing management, clinical medicine, clinical nursing, or statistics; (4) Minimum 10 years’ professional practice;(5) Willingness to participate in this study. Following the Delphi expert consultation methodology, 15 relevant experts from Jiangsu Province were selected to participate between 29 June and 31 July 2025. Their profiles are detailed in Table 1. The consultation questionnaire comprised an item evaluation scale, an expert profile survey, and an expert authority assessment form.Experts assessed the necessity of scale items and provided revision suggestions via standardised questionnaires. The expert consultation form employed a 5-point Likert scale: “Not at all important” (1 point), “Not important” (2 points), “Neutral” (3 points), “Important” (4 points), “Very important” (5 points) ^[20]^.Two rounds of expert consultation were conducted based on the initial questionnaire. The expert consultation questionnaires were distributed and collected in paper format or via email. Items retaining an importance score mean ≥4.0 and coefficient of variation ≤0.25 were retained. Items were added, removed, or modified based on expert opinions and suggestions ^[20]^.

**Table 1.** General characteristics of experts (n=15)

| Item |  | Number of Cases | Percentage |
| --- | --- | --- | --- |
| Gender | Male | 3 | 20 |
|  | Female | 12 | 80 |
| Age | 31-40 | 5 | 33.3 |
|  | 41-50 | 8 | 53.4 |
|  | Over 50 | 2 | 13.3 |
| Field of work | Clinical Medicine | 1 | 6.6 |
|  | Clinical Nursing | 8 | 53.4 |
|  | Nursing Management | 5 | 33.3 |
|  | Medical Statistics | 1 | 6.6 |
| Educational Background | Bachelor's Degree | 9 | 60 |
|  | Bachelor's degree or above | 6 | 40 |
| Professional Title | Associate Senior | 5 | 33.3 |
|  | Senior | 10 | 66.7 |

### 1.3 Preliminary Study

Preliminary Study This study was approved by the Ethics Committee of Wuxi Second People’s Hospital, with ethics approval number 2025Y-81. Using convenience sampling, 32 registered nurses from the General Intensive Care Unit (ICU) at Wuxi Second People’s Hospital were selected as subjects for the pre-test study.The resulting ’Knowledge, Attitude and Practice Scale for Oxygen Saturation Management in Mechanically Ventilated Patients’ comprises three core dimensions: Knowledge (11 items), Attitude (6 items) and Practice (9 items), totalling 26 assessment items. The scale employs a five-point Likert scale (1 = Strongly Disagree, 5 = Strongly Agree).

### 1.4 Statistical Methods

Data entry and analysis were conducted using Excel, SPSS 27.0, and AMOS 29.0 software. Item screening was performed using the Critical Ratio Method ^[20]^.Reliability was assessed by calculating Cronbach’s α and Spearman-Brown coefficients. Exploratory and confirmatory factor analyses validated the questionnaire’s construct validity, convergent validity, and discriminant validity. Content validity was evaluated using content validity indices, with a significance level of α = 0.05 ^[20]^.

## 2. Results

### 2.1. Content Validity

Between 29 June and 31 July 2025, 15 experts possessing profound professional knowledge and extensive practical experience in nursing were invited to participate. This group comprised 5 nursing management specialists, 8 ICU clinical specialists, 1 medical statistics expert, and 1 clinical medicine expert.During two consecutive rounds of expert consultation, the questionnaire response rate reached 100% in both instances, with expert authority indices exceeding 0.8. Kendall’s W consistency coefficients were 0.179 and 0.182 respectively (all P<0.05), confirming the expert group’s high engagement, authority, and consensus coherence. (^[20]^)In the first round of expert consultation, the behaviour dimension item “You select the opposite limb when a blood pressure cuff or arterial catheter is present” was revised to “You avoid placing the sensor on the same limb as a blood pressure cuff or arterial catheter”.In the knowledge dimension, the imprecise phrasing “Are you aware of the replacement frequency during continuous pulse oximetry monitoring?” was revised to “Are you aware of the replacement frequency for the site of continuous pulse oximetry monitoring?”. The second round of expert consultation yielded no further modification suggestions. Calculation of the expert consultation data yielded an overall content validity index (S-CVI) of 0.886 (Table 2), with knowledge, attitude, and practice dimensions ranging from 0.867 to 1.00. Individual item content validity indices (I-CVI) ranged from 0.913 to 1.00.

**Table 2.** Overall Content Validity Index (S-CVI)

| Item | Value | Criterion | Evaluation |
| --- | --- | --- | --- |
| S-CVI/UA | 0.886 | Value $\geq 0.8$ | Pass |
| S-CVI/Ave | 0.990 | Value $\geq 0.9$ | Pass |

\*Note: S-CVI/UA denotes the overall S-CVI, while S-CVI/Ave represents the average S-CVI

### 2.2 Item Analysis

A total of 32 questionnaires were distributed for the pre-test, with all 32 returned as valid responses. Participants were grouped by total score: the top 27% constituted the high-scoring group, and the bottom 27% the low-scoring group. An independent samples t-test was employed to compare score differences between groups ^([21]).^. The t-values for the remaining items ranged from 3.676 to 24.811, all yielding P < 0.01, indicating good discriminatory power. Differences in item scores were statistically significant (P < 0.05), and all items were retained.

### 2.3 Construct Validity

Exploratory factor analysis (EFA) was conducted using SPSS 27.0 software. Bartlett’s sphericity test revealed a KMO value of 0.918 for the total scale, with KMO values of 0.951, 0.925, and 0.949 for the knowledge, attitude, and practice dimensions respectively. The approximate chi-square value for the total scale was 4623.04, with approximate chi-square values of 1944.14, 905.6, and 141.0 for the three dimensions respectively.0.949, respectively. The approximate chi-square value for the total scale was 4623.04, while the approximate chi-square values for the three dimensions were 1944.14, 905.6, and 1416.16, respectively, all meeting statistical requirements (P<0.001). This indicates the questionnaire data is suitable for further exploratory factor analysis.Principal component analysis was employed, with maximum variance factor rotation. Common factors were extracted using eigenvalues >1.0 as the criterion ^([22]).^Ultimately, one, one, one, and three common factors were extracted for knowledge, attitude, behaviour, and the questionnaire as a whole, respectively. The knowledge dimension was named Nurses’ Overall Perception of Oxygen Saturation Management in Mechanically Ventilated Patients, the attitude dimension was named Nurses’ Willingness to

Manage Oxygen Saturation in Mechanically Ventilated Patients, and the behaviour dimension was named Nurses’ Practice of Oxygen Saturation Management in Mechanically Ventilated Patients.The cumulative variance explained by the factors was 70.402%, as detailed in Figure 1, Table 3, and Table 4.

**Table 3.**
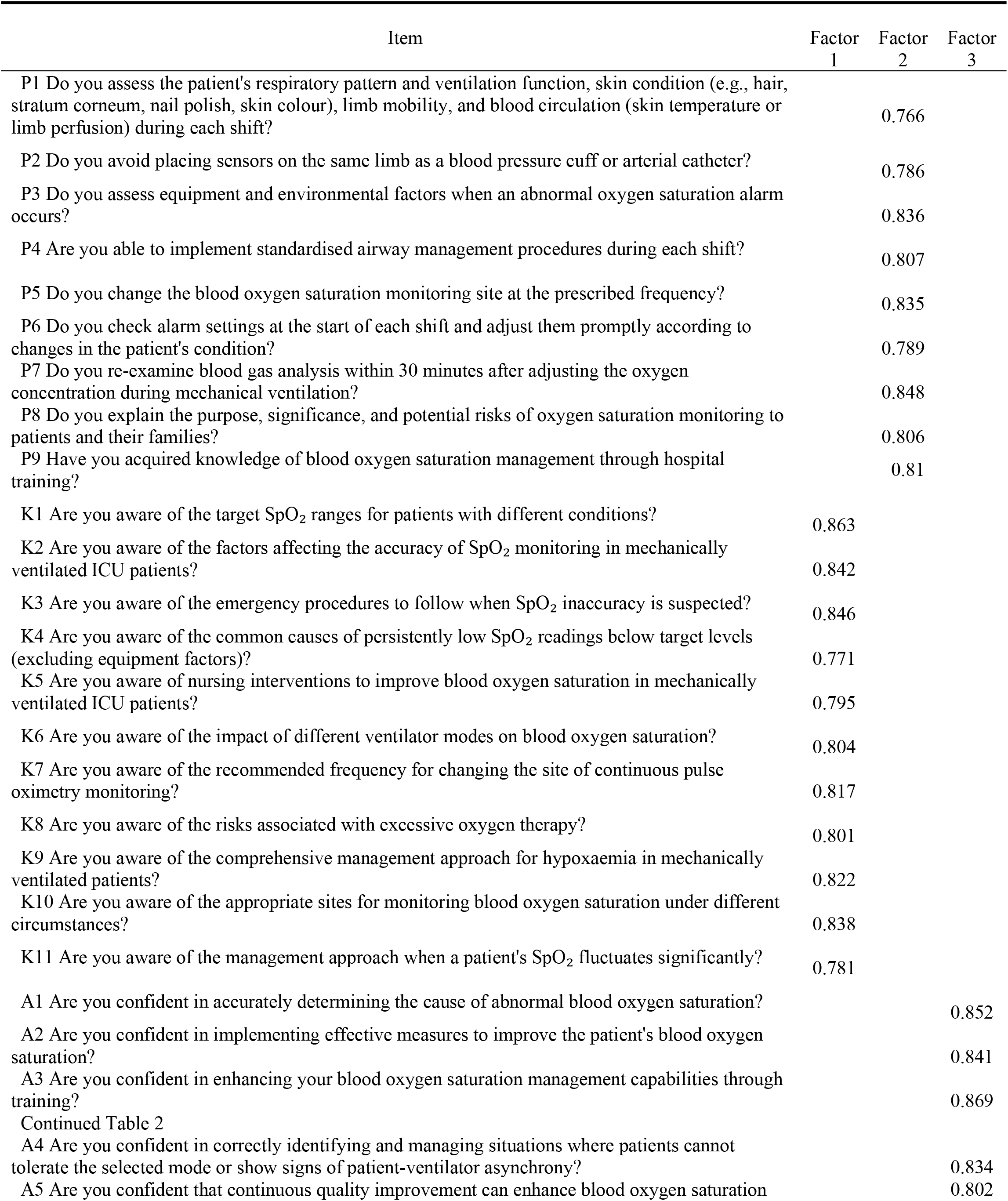
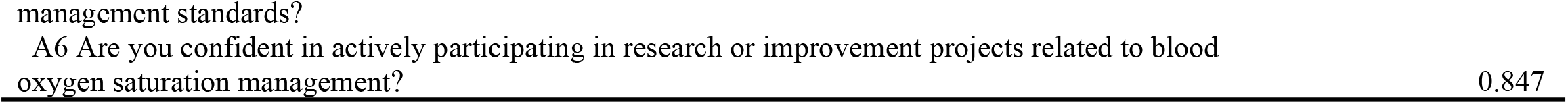
Factor analysis results of the knowledge, attitude, and practice questionnaire on blood oxygen saturation management for mechanically ventilated ICU patients among clinical nurses.

**Table 4.** Total variance explained by each factor.

| Factor | Initial Eigenvalue |  |  | Extracted load squares |  |  | Rotational loadings sum of squares |  |  |
| --- | --- | --- | --- | --- | --- | --- | --- | --- | --- |
|  | Total | Percentage of variance | Cumulative % | Grand Total | Percentage of variance | Cumulative % | Grand Total | Percentage of variance | Cumulative % |
| 1 | 10.03 | 38.575 | 38.575 | 10.03 | 38.575 | 38.575 | 7.584 | 29.168 | 29.168 |
| 2 | 4.711 | 18.119 | 56.694 | 4.711 | 18.119 | 56.694 | 6.213 | 23.897 | 53.064 |
| 3 | 3.564 | 13.707 | 70.402 | 3.564 | 13.707 | 70.402 | 4.508 | 17.337 | 70.402 |

**Fig 1.**
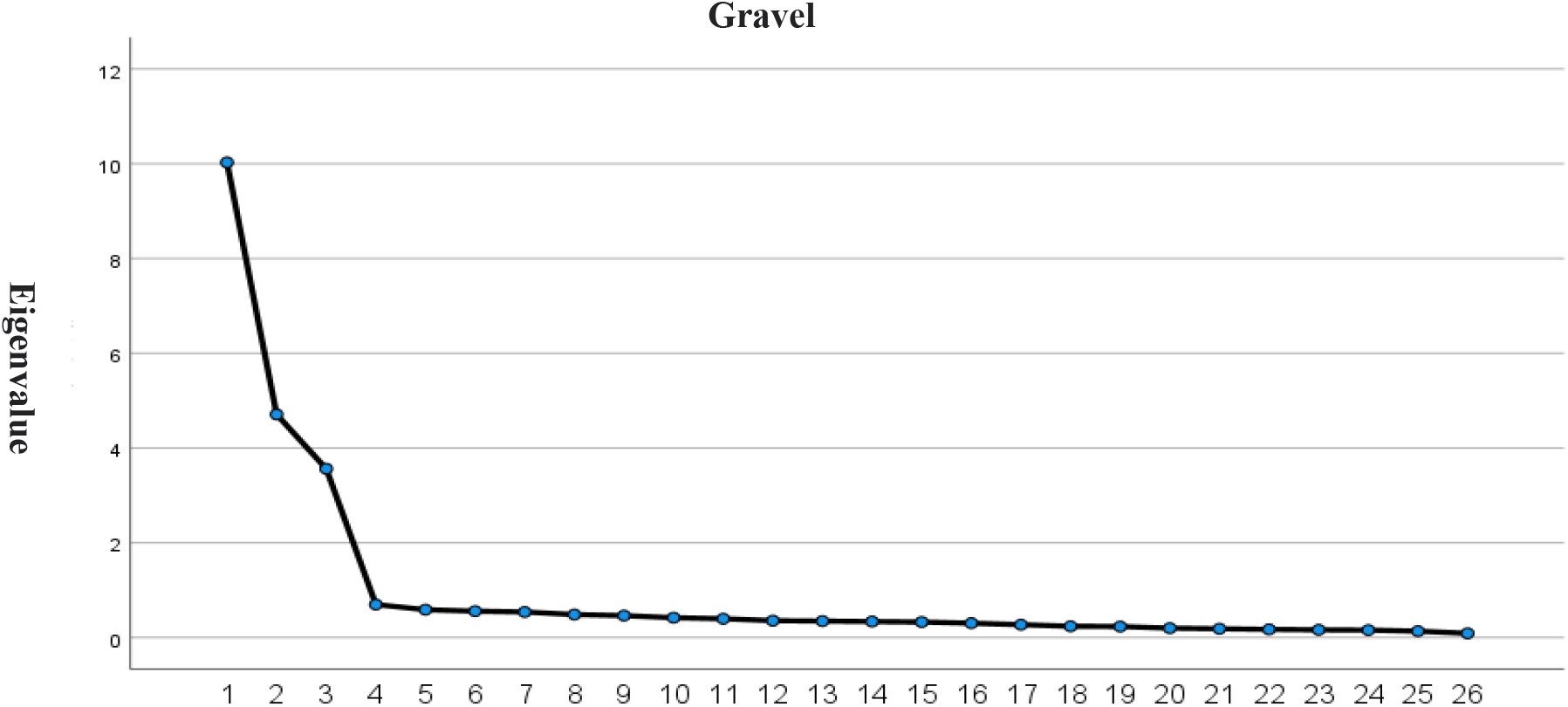
Exploratory factor analysis scatter plot of the knowledge-attitude-behaviour questionnaire on blood oxygen saturation management for mechanically ventilated patients among ICU nurses

### 2.4 Reliability analysis

Using SPSS 27.0 statistical software, Cronbach’s α coefficients were calculated for the overall questionnaire and each dimension. Cronbach’s α is a commonly used indicator for assessing internal consistency of questionnaires, with values ranging from 0 to 1. A higher coefficient indicates stronger consistency among items and greater reliability of the questionnaire.Generally, a Cronbach’s α coefficient exceeding 0.7 indicates good internal consistency ^([23]^). The Cronbach’s α coefficients for the knowledge, attitude, and behaviour dimensions were 0.687, 0.622, and 0.681 respectively, while the overall questionnaire achieved a Cronbach’s α of 0.88.The Spearman-Brown coefficients for the knowledge, attitude, and behaviour dimensions were 0.796, 0.666, and 0.728 respectively, with the overall Spearman-Brown coefficient for the questionnaire being 0.967. The test-retest reliability of this questionnaire was 0.912. Consequently, this questionnaire possesses good reliability.

### 2.5 Confirmatory factor analysis

A total of 230 questionnaires were distributed, with 219 returned, yielding a response rate of 95.2%. After excluding 7 questionnaires of poor quality, 212 valid responses were analysed. Confirmatory factor analysis (CFA) was conducted using the structural equation modelling software AMOS 29.0. A pre-specified factor model was established based on the EFA results (Figure 2), with overall model fit indices meeting basic standards (Table 5). Building upon the well-fitted CFA model, convergent and discriminant validity were further examined.As shown in Table 6, factor loadings for knowledge, attitude, and behaviour all exceeded 0.7, indicating high representativeness of items within each latent variable. Furthermore, the average variance extracted (AVE) for each latent variable exceeded 0.5, and composite reliability (CR) exceeded 0.8, confirming satisfactory convergent validity ^[23]^.Table 7 demonstrates that the absolute values of correlation coefficients are all less than the square root of the corresponding AVE, indicating that the discriminant validity of the scale is ideal ^[24]^.

**Table 5.** Fit indices for the confirmatory factor analysis model of the knowledge, attitude, and practice questionnaire for ICU nurses managing blood oxygen saturation in mechanically ventilated patients

|  | Standardised Value | Model Fit Value |
| --- | --- | --- |
| $\chi^2/df$ | <3 | 2.069 |
| Goodness-of-Fit Index (GFI) | >0.90 | 0.829 |
| Adjusted Goodness-of-Fit Index (AGFI) | >0.90 | 0.798 |
| Root Mean Square Error of Approximation (RMSEA) | <0.10 | 0.071 |
| Root Mean Square Residual (RMR) | <0.05 | 0.023 |
| Comparative Fit Index (CFI) | >0.9 | 0.930 |
| Normative Fit Index (NFI) | 0.9 | 0.874 |
| Tucker-Lewis Index (TLI) | >0.9 | 0.923 |
| Incremental Fit Index (IFI) | >0.9 | 0.930 |
| Pareto-Consistent Fitting Index (PCFI) | >0.5 | 0.847 |
| Prescriptive Normalisation Index (PNFI) | >0.5 | 0.796 |

**Table 6.**
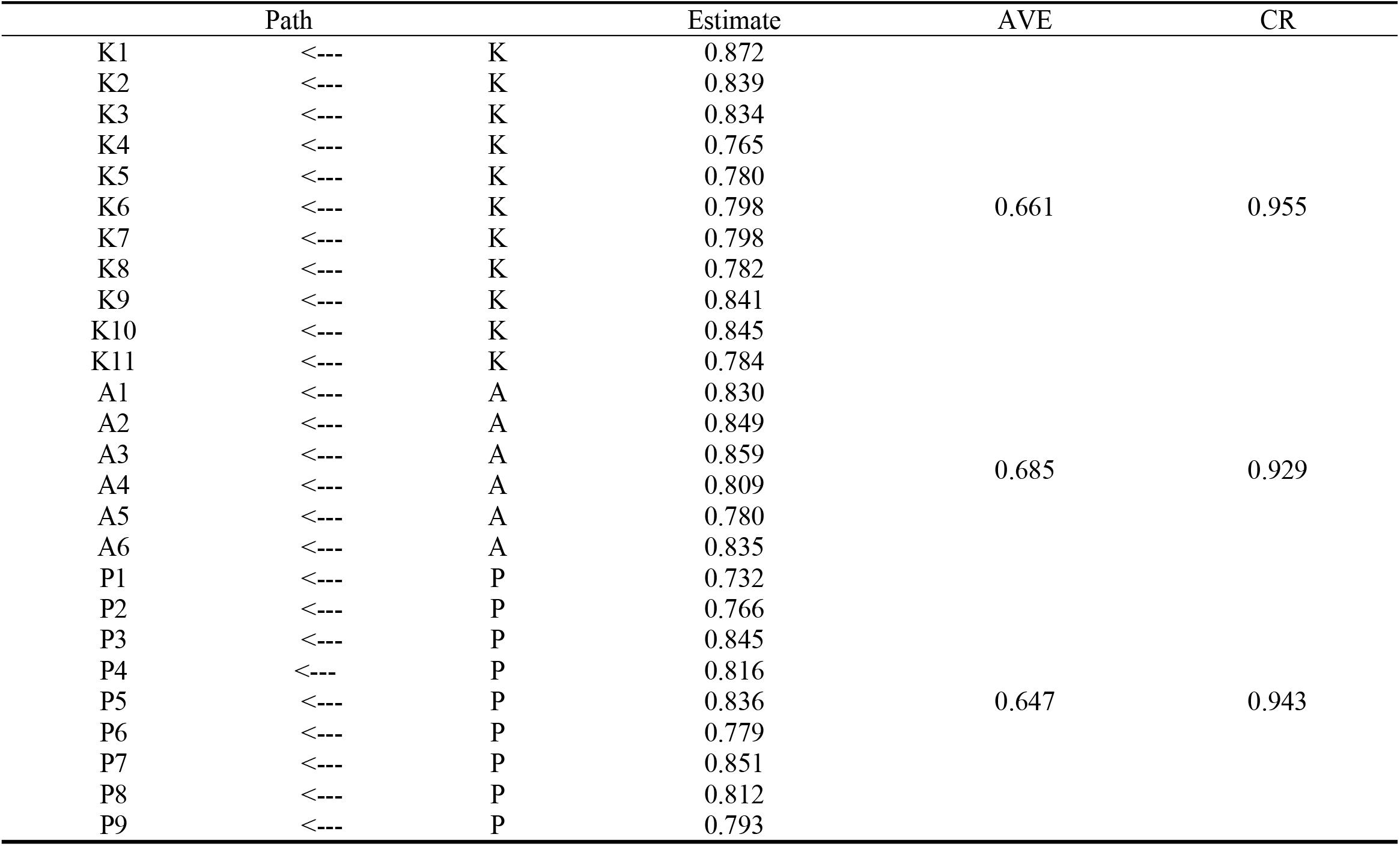
Confirmatory factor analysis of ICU nurses’ knowledge, attitude, and practice questionnaire on blood oxygen saturation management in mechanically ventilated patients Aggregate validity indices

**Table 7.** Distinctive Validity Indicators from Confirmatory Factor Analysis of ICU Nurses’ Knowledge, Attitude, and Practices Questionnaire on Oxygen Saturation Management in Mechanically Ventilated Patients

|  | K | A | P |
| --- | --- | --- | --- |
| K | 0.661 |  |  |
| A | 0.102** | 0.684 |  |
| P | 0.125** | 0.085** | 0.646 |
| AVE square root | 0.813 | 0.827 | 0.804 |

**Figure 2.**
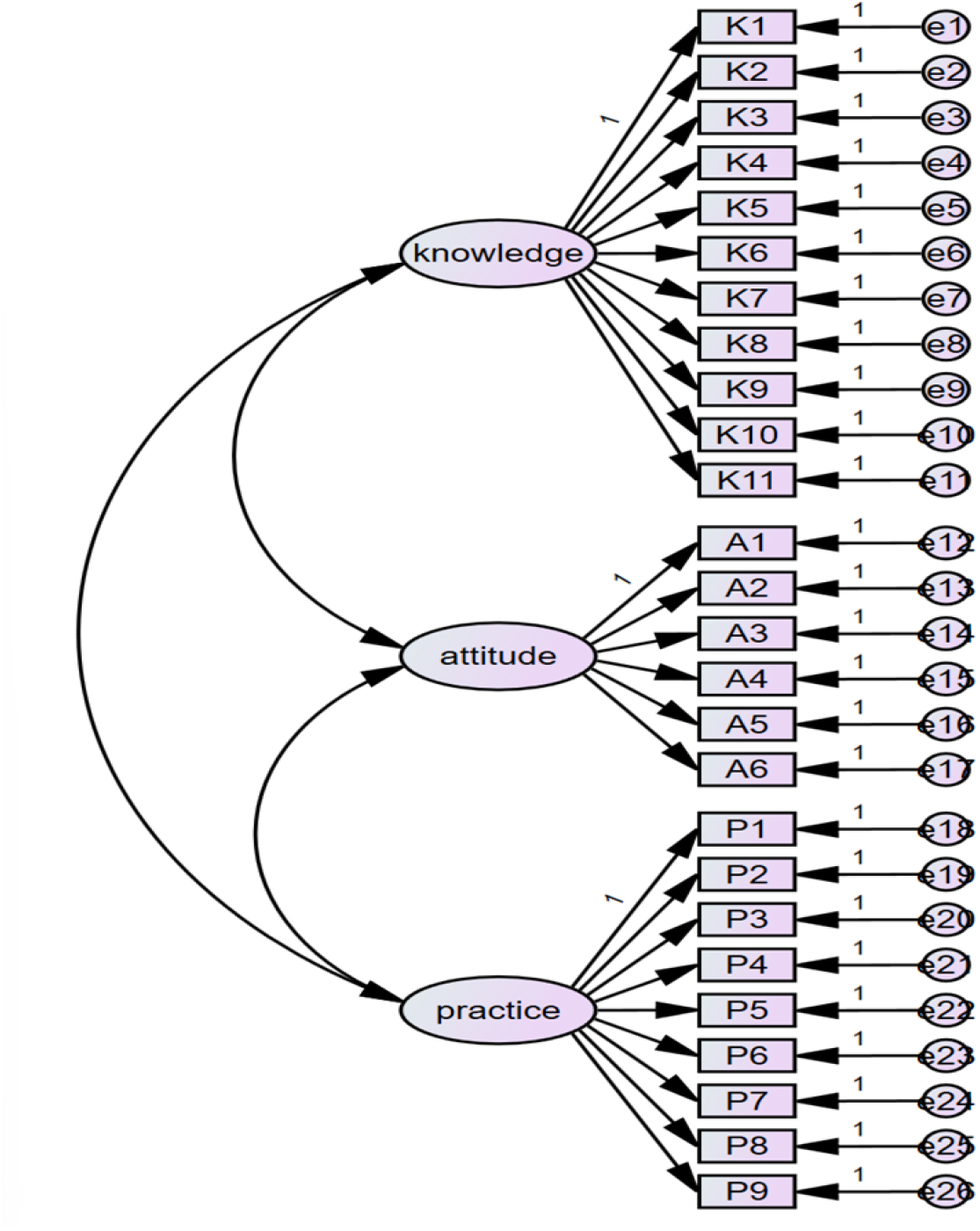
Confirmatory factor analysis model for the ICU nurses’ knowledge, attitude, and practice questionnaire on blood oxygen saturation management in mechanically ventilated patients

## 3. Discussion

### 3.1 The Knowledge-Attitude-Practice (KAP) questionnaire for ICU nurses managing SpO_2_ in mechanically ventilated patients demonstrates scientific validity and practical applicability

Grounded in KBAP theory and informed by daily ICU clinical practice, this study systematises the SpO_2_ management protocol for mechanically ventilated patients, ensuring the questionnaire comprehensively reflects clinical realities.During development, the team first reviewed relevant domestic and international literature, gathering information on fundamental principles of mechanical ventilation, SpO_2_ monitoring methods, alarm management protocols, ventilator parameter adjustments, and specific behavioural requirements for nursing interventions. This formed the basis for preliminary questionnaire items. Subsequently, two rounds of Delphi expert consultation were conducted with 15 clinical specialists possessing extensive ICU experience.Experts meticulously evaluated each item against three criteria: clinical utility, professional accuracy, and clarity of expression, subjecting them to repeated deliberation. Recent clinical advancements and practice consensus were incorporated, refining item phrasing and structural organisation to enhance scientific rigour and clinical applicability.The questionnaire was progressively refined through literature analysis, expert collaboration, and multiple revisions. The final instrument aims to systematically assess ICU nurses’ competence in managing SpO_2_ in mechanically ventilated patients across knowledge, attitude, and behaviour domains, providing guidance for clinical training and quality improvement.

### 3.2 The validity and reliability testing of the Knowledge-Attitude-Behaviour Questionnaire for ICU Nurses’ Management of SpO_2_ in Mechanically Ventilated Patients yielded favourable results

In reliability analysis, the questionnaire achieved an overall Cronbach’s α coefficient of 0.88, with all dimension coefficients falling within acceptable ranges. This indicates strong internal consistency among items, enabling stable assessment of ICU nurses’ knowledge, attitude, and practices regarding SpO_2_ management in mechanically ventilated patients.Furthermore, the split-half reliability method yielded Spearman-Brown coefficients exceeding 0.70 (P<0.01), indicating stable results across different assessment time points with minimal temporal variation. Validity was assessed through content and construct validity. Content validity was evaluated by 15 experts spanning nursing management, ICU clinical nursing, clinical medicine, and medical statistics.Results indicated item-level content validity indices (I-CVI) ranging from 0.913 to 1.00, while the scale-level content validity index (S-CVI) was 0.886. This demonstrates that the questionnaire content aligns with the knowledge, attitude, and behaviour dimensions of SpO_2_ management in mechanically ventilated patients, effectively covering key elements in this field. Construct validity was supported by exploratory factor analysis and confirmatory factor analysis.Exploratory factor analysis yielded three common factors representing knowledge, attitude, and behaviour dimensions respectively. The cumulative variance explained met requirements, and factor loadings for all items fell within acceptable ranges, confirming the questionnaire’s clear structure and ability to distinguish between dimensions.The confirmatory factor analysis results demonstrated overall favourable model fit indices, with all fit measures within desirable ranges and factor loadings for each item falling within reasonable intervals. This provides empirical support for the questionnaire structure, indicating that the theoretical model effectively reflects the current state of knowledge, attitude, and practice concerning SpO_2_ management among ICU nurses in their clinical work.

### 3.3 The questionnaire on ICU nurses’ knowledge, attitude, and practices regarding SpO_2_ management in mechanically ventilated patients provides scientific grounds for developing personalised training programmes

This questionnaire holds considerable practical value in ICU clinical practice. It can be used to systematically evaluate nurses’ knowledge, attitude, and behaviours concerning SpO_2_ management in mechanically ventilated patients, identifying specific weaknesses in this area and thereby informing targeted training initiatives.Based on assessment outcomes, diverse training formats—such as thematic lectures, clinical case discussions, and scenario simulations—can be designed to progressively enhance nurses’ comprehensive competencies in this domain. Additionally, this questionnaire serves as an evaluation tool for assessing the efficacy of subsequent training or interventions.Comparing questionnaire scores before and after intervention provides a tangible measure of improvements in nurses’ knowledge, attitude, and practice. Employing a prospective study design to track patient outcomes—including SpO_2_ target attainment rates, complication incidence, mechanical ventilation duration, and length of hospital stay—enables a more comprehensive evaluation of the nursing intervention’s practical efficacy. This provides evidence-based grounds for refining clinical nursing strategies, thereby advancing nursing quality improvement and facilitating the clinical application of research findings.

## 4 Conclusions

This study developed a questionnaire assessing ICU nurses’ knowledge, attitude, and practices regarding SpO_2_ management in mechanically ventilated patients, and validated its reliability and validity. Results indicate the questionnaire possesses good reliability and validity, enabling stable and accurate measurement of ICU nurses’ knowledge, attitude, and behavioural levels concerning SpO_2_ management in mechanically ventilated patients.Regarding reliability, all metrics met high standards, ensuring the reliability and stability of the questionnaire’s measurement outcomes. For validity, content validity was endorsed by multidisciplinary expert review, while construct validity was supported by factor analysis, indicating that the questionnaire’s dimensional structure aligns closely with clinical practice.The study sample primarily comprised nurses from tertiary hospitals within the same region, lacking sufficient representation from nurses across different regions or ICU tiers. Given variations in patient condition profiles, nursing protocols, and training systems across institutions, adaptation to specific contexts is necessary when implementing the questionnaire.The questionnaire survey may also be influenced by social desirability bias, with some nurses’ responses not fully reflecting actual circumstances, thereby introducing potential bias in the results. Future research could increase the sample size and incorporate cross-contextual validity assessments of the questionnaire among ICU nurses from different levels and regions. Concurrently, qualitative interviews or observational methods could be employed to contextualise and revise questionnaire items, thereby enhancing its practicality across diverse clinical settings.

## Data Availability

All data generated from the custom-designed questionnaire, as well as a full copy of the questionnaire itself, have been submitted as supplementary materials with this manuscript. The corresponding author can be contacted for additional information regarding the data.

## Author contributions

Conceptualization: Hong Bian

Writing – original draft: Qian Xie, Hong Bian

Writing – review & editing: Qian Xie, Hong Bian,Ping Yu,Ping He

## References

1. Hunter S, Manias E, Hirth S, et al. Intensive care patients receiving vasoactive medications:a retrospective cohort study. Australian Critical Care. 2022;35(5):499–505.

2. Gomes EP, Reboredo MM, Costa GB, et al. Impacts of a fraction of inspired oxygen adjustment protocol in COVID-19 patients under mechanical ventilation:a prospective cohort study. Medicina Intensiva(English Edition). 2023;47(4):212–220.

3. Elhabashy S, Moriyama M, Mahmoud EIED, et al. Effect of evidence-based nursing practices training programme on the competency of nurses caring for mechanically ventilated patients:a randomised controlled trial. BMC Nursing. 2024;23(1).10.1186/s12912-024-01000-x

4. Li B, Zhang Y, Yan MQ, et al. Development of a knowledge-attitude-behaviour questionnaire for emergency department nurses regarding emerging infectious diseases. Journal of Fudan University(Medical Edition). 2023;50(1):114–121.

5. Lin WX. A study on influencing factors and prevention strategies for neck and low back pain among military flight personnel based on the knowledge-attitude-behaviour theory. 2022. 10.27002/d.cnki.gsjyu.2022.000004

6. UpToDate. Overview of initiating invasive mechanical ventilation in adults in the intensive care unit. 2025. Availablefrom: http://www-uptodate-cn-s.zju.sjuku.top/contents/zh-Hans/overview-of-initiating-invasive-mechanical-ventilation-in-adults-in-the-intensive-care-unit7.

7. UpToDate. Pulse. oximetry. 2025. Availablefrom: http://www-uptodate-cn-s.zju.sjuku.top/contents/zh-Hans/pulse-oximetry

8. Ahmad I, El-Boghdadly K, Bhagrath R, et al. Difficult Airway Society guidelines for awake tracheal intubation (ATI) in adults.1 Anaesthesia. 2020;75(4):509–528. 10.1111/anae.14904

9. Evans L, Rhodes A, Alhazzani W, et al. Surviving Sepsis Campaign: international guidelines for management of sepsis and septic shock 2021. Critical Care Medicine. 2021;49(11):e1063–e1143.10.1097/CCM.0000000000005337

10. Del Sorbo L, Goligher EC, McAuley DF, et al. Mechanical ventilation in adults with acute respiratory distress syndrome: summary of the experimental evidence for the clinical practice guideline. Annals of the American Thoracic Society. 2017;14(Suppl 4):S261–S270. 10.1513/AnnalsATS.201704-345OT

11. Siemieniuk RAC, Chu DK, Kim LH, et al. Oxygen therapy for acutely ill medical patients: a clinical practice guideline. BMJ. 2018;363:k4169. 10.1136/bmj.k4169

12. Li L, Zhang Y, Wang P, et al. Conservative versus liberal oxygen therapy for acutely ill medical patients: a systematic review and meta-analysis. International Journal of Nursing Studies. 2021;118:103924. 10.1016/j.ijnurstu.2021.103924

13. Crescioli E, Krejberg KU, Klitgaard TL, et al. Long-term effects of lower versus higher oxygenation levels in adult ICU patients—a systematic review. Acta Anaesthesiologica Scandinavica. 2022;66(8):910–922. 10.1111/aas.14107

14. Abdelbaky AM, Elmasry WG, Awad AH. Lower versus higher oxygenation targets for critically ill patients: a systematic review. Cureus. 2023;15(7):e41330. 10.7759/cureus.41330

15. Chinese Research Hospital Association. T/CRHA089—2024: nursing protocol for bedside electrocardiographic monitoring in adults. 2024.

16. Lu HL, Wang HH, Hua JN, et al. Evidence-based summary of conservative oxygen therapy nursing for adult critically ill patients. Chinese Journal of Critical Care Nursing. 2025;6(6):751–757.

17. Zhang Y, Chen XP, Shao S, et al. Evidence-based summary of alarm management for bedside electrocardiographic monitors. Chinese Journal of Nursing. 2021;56(3):445–451.

18. Managing alarms in acute care across the life span: electrocardiography and pulse oximetry. Critical Care Nurse. 2018;38(2):e16–e20. 10.4037/ccn2018468

19. Expert Consensus Group on Emergency Oxygen Therapy. Expert consensus on emergency oxygen therapy. Chinese Journal of Emergency Medicine. 2018;27(4):355–360.

20. Xu YF, Shang LP, Pan W, et al. Development and validity testing of a knowledge-attitude-practice questionnaire for clinical nurses’ participation in antimicrobial stewardship. Nursing Research. 2021;35(11):1925–1930.

21. Zhu JR, Gao MM, Jia CX, et al. Development and validity testing of a knowledge, attitudes, and practices questionnaire on blood phosphorus control in haemodialysis patients. Nursing Research. 2024;38(15):2662–2667.

22. Sun J, Yang ZL, Fan XG, et al. Development and validity testing of a knowledge-attitude-behaviour scale for disaster nursing. Evidence-Based Nursing. 2022;8(2):228–231.

23. Chen LH, Wang YN, Xing YR, et al. Development and validity testing of the knowledge, attitudes and practices scale for volume management in patients with chronic heart failure. Evidence-Based Nursing. 2025;11(22):4658–4663.

24. Wu ML. Structural equation modelling: operation and application of AMOS. Chongqing: Chongqing University Press; 2010.

